# Effect of the *INSR* gene variants on the long-term response to a 31-month childhood obesity intervention

**DOI:** 10.1101/2022.05.26.22275619

**Authors:** Jing Chen, Rui Shan, Wu-Cai Xiao, Zheng Liu

## Abstract

**Objective:** To explore effects of the *INSR* genotype on the waist circumference reduction after a lifestyle-based obesity intervention.

**Methods:** This was a nested study in a cluster-randomized controlled trial conducted from September 2018 to June 2019 in Beijing, China. Four schools (200 children) were randomized to the intervention group (diet and physical activity) and 4 schools (193 children) were randomized to the control group (usual practice without a focus on obesity prevention). We followed up children at 9 months (the end of the intervention) and 31 months (22 months after the intervention), and genotyped 7 independent SNPs in the *INSR* gene. We assessed genetic effects on changes in five waist-related indicators [waist circumference, waist-to-hip ratio (whr), waist-to-height ratio (WHtR), waist circumference adjusted by BMI (WCadjBMI), waist-to-hip ratio adjusted by BMI (WHRadjBMI)] from baseline to 9 months and from 9 months to 31 months in the intervention and control group, respectively, and compared whether genetic effects differed by group (i.e., gene-group interaction).

**Results:** From baseline to 9 months, we found that *INSR* rs7508679, rs10420008, rs11883325, and rs4804416 modified the intervention effects on changes in all waist-related indicators (all *P* < 0.05). In the control group, the effect allele was associated with greater increases in waist-related indicators, whereas opposite-directional associations were observed in the intervention group. Such interactions between SNPs and group assignment were almost not observed from 9 months to 31 months.

**Conclusion:** Our data suggested that children carrying effect alleles of rs7508679, rs10420008, rs11883325, or rs4804416 may benefit more from a lifestyle intervention for obesity prevention, but the effect appeared to be attenuated in the long term.

## INTRODUCTION

Childhood obesity has become a serious public health issue in China and worldwide. From 1975 to 2016, the prevalence of childhood obesity has increased 8 times globally^[1]^, and remains rising in low- and middle-income countries such as China^[2]^. Obesity not only influences physical and psychological health during childhood^[3]^, but also increases the risk of cardiometabolic diseases, musculoskeletal problems, and cancer in the long term^[4]^.

Effective and timely interventions are urgently needed to prevent childhood obesity. Notably, individual responses to the same intervention differ by genetic background^[5]^. Taken evidence from monozygotic twins as an example, the extent of weight loss induced by a very-low-calorie-diet intervention was very similar among intra-pair individuals (with similar genetic backgrounds); by contrast, a 12.8 times larger variability in weight loss was observed in inter-pair individuals (with different genetic backgrounds)^[6]^. Therefore, it is critical to clarify genetic effects on individuals’ responses to childhood obesity interventions.

Until now, researchers have explored the moderating effects of several glucose-metabolism-related genes on childhood obesity interventions, including *TCF7L2*^[7]^, *Visfatin*^[8]^, *MTNR1B*^[9]^, *RPTOR*^[10]^, and *SH2B1*^[11]^. Nevertheless, none of these genes have shown evidence of influences on the effects of childhood obesity interventions except for *RPTOR*^[10]^. Moreover, these genes have been examined in single-group, pre-post studies (without comparison group) and the interventions lasted for no more than a year. *INSR*, another glucose-metabolism-related gene, was associated with obesity according to Genome-Wide Association Studies (GWAS)^[12]^. But to the best of our knowledge, only one European randomized controlled trial has explored the influence of *INSR* gene variants on the effect of a 24-week obesity intervention in 445 adult population who were obese^[13]^. It remains unclear whether *INSR* is responsible for differential obesity prevention after an intervention among the children sample.

To fill the research gaps, we conducted a rigorously-designed, prospective, parallel-group randomized trial to systematically explore the influence of the *INSR* gene on the response to lifestyle-based obesity intervention in Chinese children aged 8-10 years. We first studied the influence of *INSR* gene on multiple obesity-related indicators in the intervention and control arms respectively and examined whether genetic effects differed by arm (i.e., the gene-arm interaction). Specifically, we focused on the changes of obesity-related indicators during a follow-up period as long as 31 months (including a 9-month intervention period and a 22-month follow-up period after the end of the intervention). We also examined the interactions of *INSR* gene variants and specific diet and physical activity behaviors, to clarify the specific intervention components potentially responsible for interacting with the genetic effects.

## METHODS

### Study context

The present study is nested in a cluster-randomized controlled trial of a multifaceted intervention for prevention of obesity in primary school children, named DECIDE-Children, which has been elaborated^[14]^ and prospectively registered at ClinicalTrials.gov (NCT03665857). Briefly, the DECIDE-Children project was conducted in eastern (Beijing), central (Changzhi City, Shanxi Province), and western (Urumqi City, Xinjiang Province) China from September 2018 (baseline investigation) to June 2019 (9-month follow-up, end of the intervention), and the last follow-up survey was completed 22 months after the end of intervention (31-month follow-up). A total of 24 schools and 1392 children were included in the DECIDE-Children project. The intervention successfully reduced children’s body mass index (BMI) and other adiposity outcomes (e.g., BMI Z-score, body fat percentage, waist circumference, the prevalence of obesity)^[14]^.

### The present study

For practical considerations, the present study recruited children in 8 primary schools in Beijing from the DECIDE-Children project. Except for 36 individuals who were lost to follow up, we obtained the informed written consent from the remaining 456 individuals for the collection of saliva DNA samples, 89% (407) of which agreed to participate in the present study. To be eligible for this present study, DNA samples from the study subjects should pass the quality control (described in the Genotyping below), and the values of BMI at baseline and 9-month follow-up were available. Finally, a total of 382 children were included for the present study. This study was ethically approved by the Peking University Institution Review Board (IRB00001052-20058).

### Obesity-related indicators

Children underwent standard physical examination including measurements of height, body weight, waist circumference, hip circumference, and body fat percentage^[14]^ at baseline, 9 months (the end of the intervention), and 31 months (22 months after the end of the intervention). Obesity-related indicators were categorized into BMI-, waist-related indicators, and body fat percentage. BMI-related indicators included body weight, BMI [calculated as body weight (kg) / height (m)^2^], and BMI Z-score^[15]^. Waist-related indicators included waist circumference, waist-to-hip ratio (whr), waist-to-height ratio (WHtR), waist circumference adjusted for BMI (WCadjBMI), and waist-to-hip ratio adjusted for BMI (WHRadjBMI). See details of waist-related indicators in the Supplemental Material.

### Genotyping

We extracted DNA from saliva samples using the buccal swab genomic DNA extraction kit (spin column type). Genetic polymorphisms were detected by a high-throughput genotyping chip (Illumina ASA) specially designed for Asians. Quality control criteria of DNA samples included SNP-level missing < 2%, individual-level missing < 2%, minor allele frequency (MAF) > 5%, Hardy-Weinberg equilibrium p-value > 1e-6, heterozygosity rate < 3 standard deviations, and no relatedness^[16]^.

We selected 7 independent SNPs (linkage disequilibrium measured by r^2^ < 0.80) meeting the quality control criteria in the *INSR* gene, including rs7508679 (MAF = 0.490), rs2115386 (MAF = 0.437), rs10420008 (MAF = 0.417), rs7248939 (MAF = 0.407), rs11671297 (MAF = 0.402), rs11883325 (MAF = 0.392), and rs4804416 (MAF = 0.391) (Figure 1).

**Figure 1.**
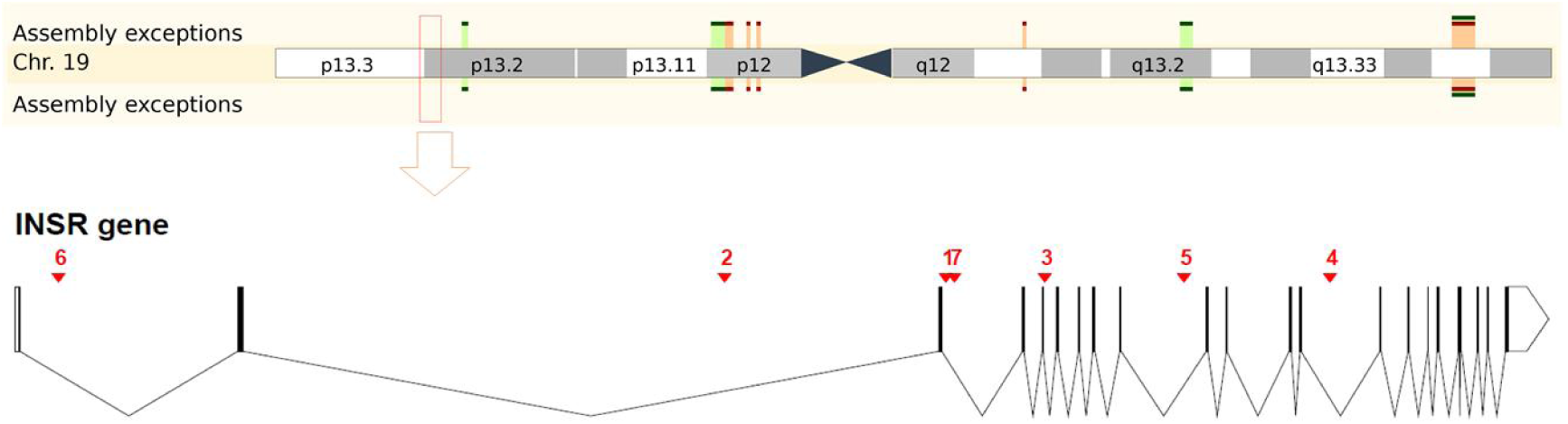
The upper half is a figure of chromosome 19 adapted from Ensembl portal (https://grch37.ensembl.org/Homo_sapiens/Location/View?r=19:7112265-7294414). The lower half is a concise genomic map for 7 SNPs within the *INSR* gene. The location of the SNPs is plotted along with the same genomic coordinates at the bottom. SNP IDs 1-7 refer to rs7508679, rs2115386, rs10420008, rs7248939, rs11671297, rs11883325, and rs4804416, respectively.

**Figure 2.**
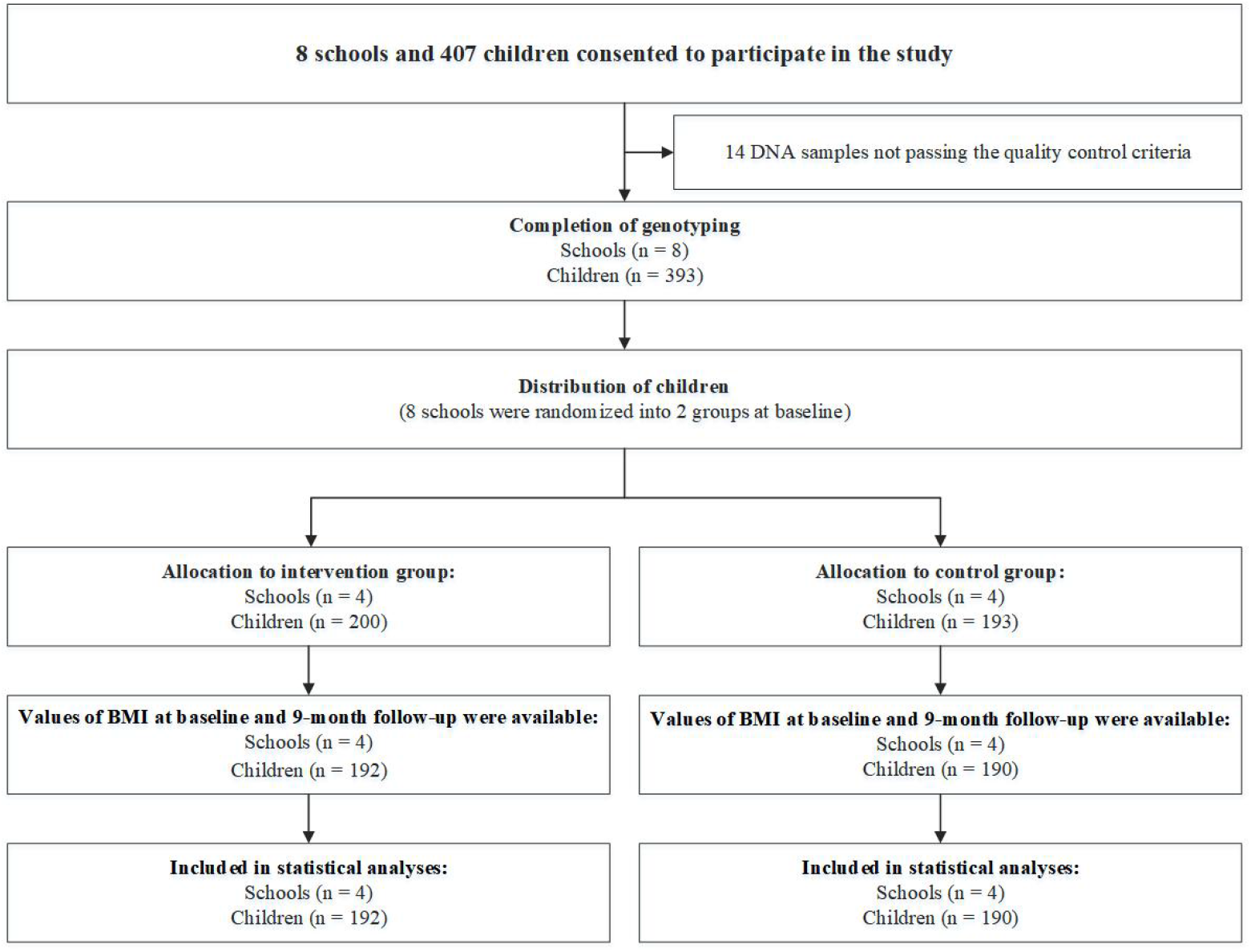
Study flow

### Statistical analyses

First, we examined cross-sectional associations between *INSR* gene variants and obesity-related indicators at baseline with adjustment for sex and age. Then, we assessed the gene-group interactions on changes in obesity-related indicators from baseline to 9 months. To test for interactions, we fitted the multiple linear regression model for each SNP: obesity-related indicators at 9 months = β_0_ + β_1_ * group + β_2_ * genotype + β_3_ * sex + β_4_ * age + β_5_ *obesity-related indicators at baseline + β_6_* genotype × group. Next, we examined the gene-group interactions on changes in obesity-related indicators from 9 months to 31 months by using the model for each SNP: obesity-related indicators at 31 months = β_0_ + β_1_ * group + β_2_ * genotype + β_3_ * sex + β_4>_ * age + β_5_ *obesity-related indicators at 9 months + β_6_* genotype × group. Lastly, we assessed gene-behavior interactions on obesity-related indicators at baseline. See details of variables of diet and physical activity behaviours in the Supplemental Material. We used additive genetic model in the main analyses, and used dominant genetic model in the sensitivity analyses. To test the robustness of study findings, we also focused on a sample of children who were overweight or obese in the sensitivity analyses. We used Quanto 1.2.4 to estimate the detectable effect sizes of gene-group interactions. The study had 80% power to detect gene-arm interaction effect size of 0.34 units for changes in WHRadjBMI at 9 months at a significance level of 0.05. Statistical analyses were performed with R 4.1.0.

## Results

### Characteristics of the study population according to genotype

Baseline characteristics of participants according to distribution of genotype are presented in Table 1 (*INSR* rs4804416) and Supplemental Tables 1-6 (other SNPs in *INSR*). There was no difference in the distribution of genotypes by sex or group. Most obesity-related indicators (body weight, BMI, BMI Z-score, waist circumference, whr, WHtR, and body fat percentage) were not related to the distribution of *INSR* 7 SNPs’ genotypes at baseline, except that WCadjBMI and WHRadjBMI were related to the distribution of rs2115386 genotype.

**Table 1.**
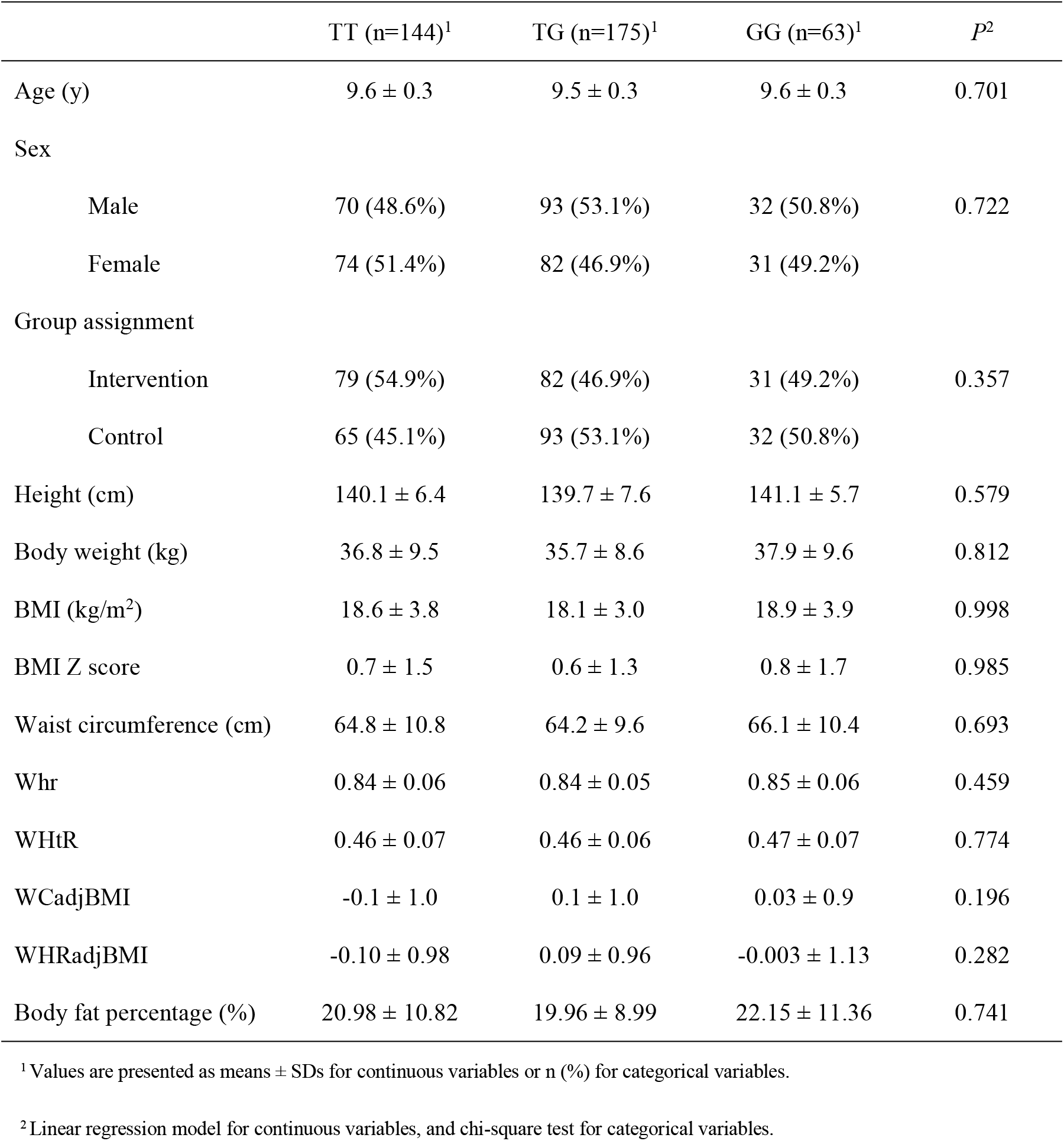
Baseline characteristics of the study population according to *INSR* rs4804416 genotype.

|  | TT (n=144)<sup>1</sup> | TG (n=175)<sup>1</sup> | GG (n=63)<sup>1</sup> | *P*<sup>2</sup> |
| --- | --- | --- | --- | --- |
| Age (y) | 9.6 ± 0.3 | 9.5 ± 0.3 | 9.6 ± 0.3 | 0.701 |
| Sex |  |  |  |  |
| Male | 70 (48.6%) | 93 (53.1%) | 32 (50.8%) | 0.722 |
| Female | 74 (51.4%) | 82 (46.9%) | 31 (49.2%) |  |
| Group assignment |  |  |  |  |
| Intervention | 79 (54.9%) | 82 (46.9%) | 31 (49.2%) | 0.357 |
| Control | 65 (45.1%) | 93 (53.1%) | 32 (50.8%) |  |
| Height (cm) | 140.1 ± 6.4 | 139.7 ± 7.6 | 141.1 ± 5.7 | 0.579 |
| Body weight (kg) | 36.8 ± 9.5 | 35.7 ± 8.6 | 37.9 ± 9.6 | 0.812 |
| BMI (kg/m<sup>2</sup>) | 18.6 ± 3.8 | 18.1 ± 3.0 | 18.9 ± 3.9 | 0.998 |
| BMI Z score | 0.7 ± 1.5 | 0.6 ± 1.3 | 0.8 ± 1.7 | 0.985 |
| Waist circumference (cm) | 64.8 ± 10.8 | 64.2 ± 9.6 | 66.1 ± 10.4 | 0.693 |
| W<sub>hr</sub> | 0.84 ± 0.06 | 0.84 ± 0.05 | 0.85 ± 0.06 | 0.459 |
| W<sub>HtR</sub> | 0.46 ± 0.07 | 0.46 ± 0.06 | 0.47 ± 0.07 | 0.774 |
| W<sub>C</sub>adjBMI | -0.1 ± 1.0 | 0.1 ± 1.0 | 0.03 ± 0.9 | 0.196 |
| W<sub>H</sub>RadjBMI | -0.10 ± 0.98 | 0.09 ± 0.96 | -0.003 ± 1.13 | 0.282 |
| Body fat percentage (%) | 20.98 ± 10.82 | 19.96 ± 8.99 | 22.15 ± 11.36 | 0.741 |

<sup>1</sup> Values are presented as means ± SDs for continuous variables or n (%) for categorical variables.
<sup>2</sup> Linear regression model for continuous variables, and chi-square test for categorical variables.

### Gene-group interactions on changes in obesity-related indicators from baseline to 9 months

#### Gene-group interactions on changes in waist-related indicators

Table 2 reports the statistically significant interactions between genotype and group assignment on changes in waist-related indicators. *INSR* rs7508679, rs10420008, rs11883325, and rs4804416 significantly modified intervention effects on waist, whr, WHtR, WCadjBMI, or WHRadjBMI at 9 months (all *P* < 0.05).

**Table 2.**
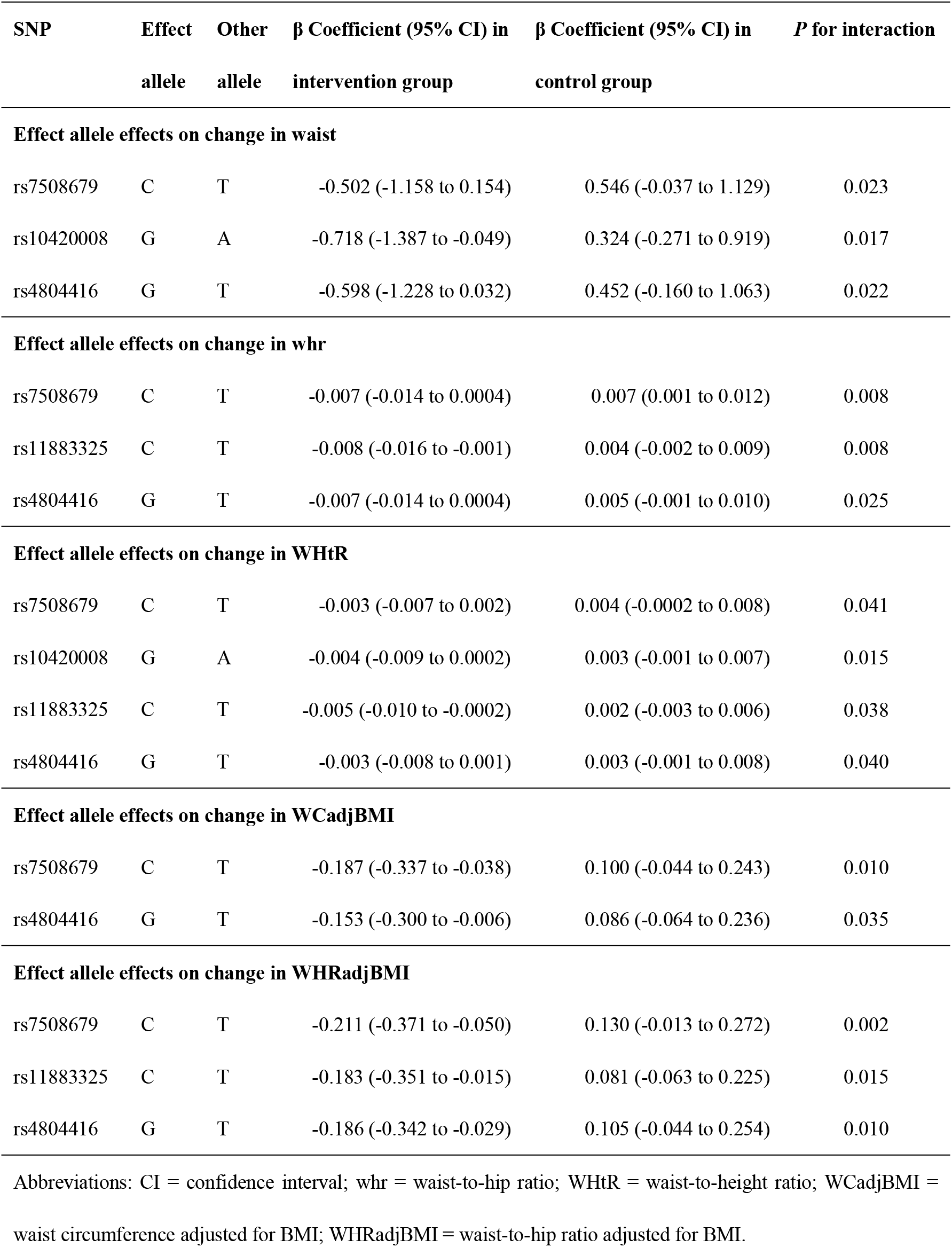
Statistically significant moderation by *INSR* genotype of intervention effects on change in waist, whr, WHtR, WCadjBMI, and WHRadjBMI.

#### Gene-group interactions on changes in body weight, BMI, and BMI Z-score

Table 3 reports the statistically significant interactions between genotype and group assignment on changes in body weight, BMI, and BMI Z-score. Only *INSR* rs10420008 significantly modified intervention effects on body weight, BMI, and BMI Z-score at 9 months (all *P* < 0.05).

**Table 3.**
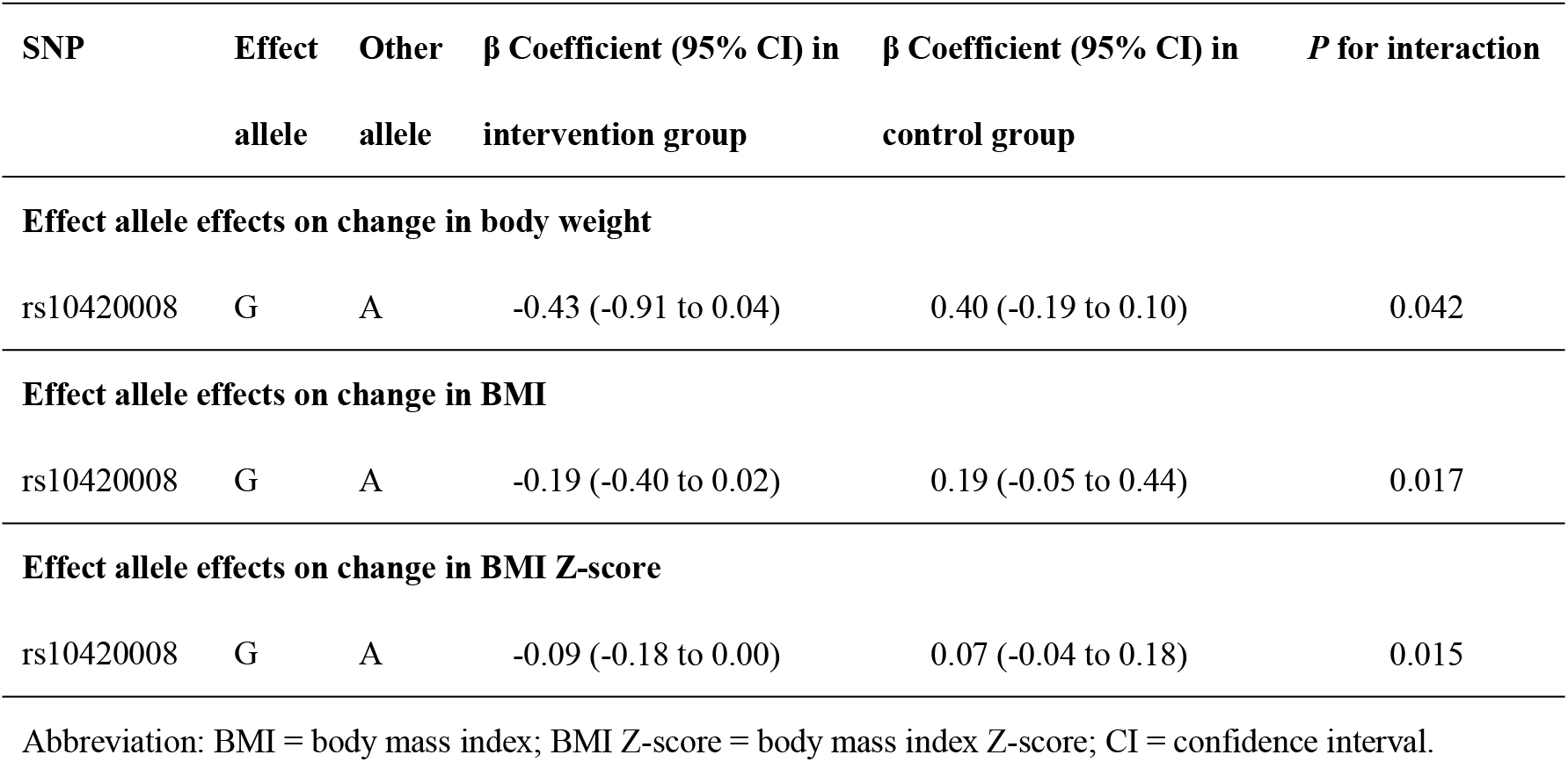
Statistically significant moderation by *INSR* genotype of intervention effects on change in body weight, BMI, and BMI Z-score.

| SNP | Effect allele | Other allele | $\beta$ Coefficient (95% CI) in intervention group | $\beta$ Coefficient (95% CI) in control group | *P* for interaction |
| --- | --- | --- | --- | --- | --- |
| **Effect allele effects on change in body weight** |  |  |  |  |  |
| rs10420008 | G | A | -0.43 (-0.91 to 0.04) | 0.40 (-0.19 to 0.10) | 0.042 |
| **Effect allele effects on change in BMI** |  |  |  |  |  |
| rs10420008 | G | A | -0.19 (-0.40 to 0.02) | 0.19 (-0.05 to 0.44) | 0.017 |
| **Effect allele effects on change in BMI Z-score** |  |  |  |  |  |
| rs10420008 | G | A | -0.09 (-0.18 to 0.00) | 0.07 (-0.04 to 0.18) | 0.015 |

Abbreviation: BMI = body mass index; BMI Z-score = body mass index Z-score; CI = confidence interval.

We didn’t find any statistically significant interaction between 7 SNPs and group assignment on changes in body fat percentage during the intervention. The gene-group interactions on obesity-related indicators among all 7 SNPs are presented in Supplemental Tables 7-9.

In the sensitivity analyses, we observed that neither a dominant model nor narrowing the study participants to those overweight or obese could largely influence the results from the main analyses (Supplemental Tables 10-15).

### Gene-group interactions on the changes of obesity-related indicators from 9 months to 31 months

From 9 months to 31 months, we didn’t find any gene-group interaction on the changes of obesity-related indicators (Supplemental Tables 16-17), except that *INSR* rs2115386 interacted with group assignment on changes in BMI (*P* = 0.039).

### Gene-behavior interactions on the obesity-related indicators

Interestingly, the specific composition of behaviors that could modify genetic effects on the levels of obesity-related indicators varied by SNP. We found *INSR* rs7508679, rs10420008, rs7248939, and rs11883325 interacted with both diet and physical activity behaviors, while rs11671297 and rs4804416 interacted with only diet behaviors with regard to obesity-related indicators at baseline (Supplemental File 1). No interactions were found between rs2115386 and behaviors on any of the indicators.

## Discussion

In this rigorously-designed, parallel-group randomized trial, we found that *INSR* rs7508679, rs10420008, rs11883325, and rs4804416 interacted with the group assignment on the changes in waist-related indicators from baseline to the end of the intervention (9 months). In the intervention group, the effect allele was associated with greater decreases in waist-related indicators, whereas in the control group opposite-directional associations were observed. Such effects were almost not observed on BMI indicators. And genetic effects on the response to the intervention did not sustain to 31 months.

Previous studies have explored genetic effects on the response to childhood obesity interventions. For example, a single-group trial^[10]^ implemented a hospital-based lifestyle intervention among German children who were overweight or obese, and discovered 9 of 56 obesity SNPs (e.g. *RPTOR* rs12940622, *CADM2* rs13078960, etc.) correlated with the changes of body weight, BMI or BMI-z scores; another one-year lifestyle intervention^[7]^ among 236 German children who were overweight or obese have indicated that *TCF7L2* rs7903146 was related to glucose metabolism after the intervention; findings from a four-week aerobic exercise training program^[8]^ in 88 Chinese children who were overweight or obese showed that *Visfatin* variant rs4730153 was associated with changes of Homeostasis model assessment of β-cell function after the program. Findings from these studies suggested that genetics factors, especially glucose-metabolism-related genes, play a critical role in childhood obesity interventions; however, there has been virtually no study based on randomized controlled trials.

As far as we know, we have made the first attempt to explore effects of *INSR* on the response to the childhood obesity intervention based on a 31-month, randomized, parallel-group controlled trial. Several lines of evidence suggest that the observed interactions between the *INSR* genotypes and childhood obesity intervention are biologically plausible. For example, an obesity intervention ^[13]^ among adults with obesity has revealed that *INSR* rs4804428, rs2396185, and rs10419421 were interacted with the drug treatment on changes in weight after the intervention. Additionally, other genes in the insulin signaling pathway were shown to modify the effect of childhood obesity interventions as *INSR* did. For example, Heitkamp^[10]^ et al. implemented a lifestyle intervention in 1198 German children with obesity and found that the A allele of *RPTOR* rs12940622 (in the insulin signaling pathway) was associated with greater weight loss after the intervention. Evidence from animal studies^[17]^ and observational studies^[18]^ have showed that *INSR* variants were associated with insulin resistance or decreased insulin sensitivity, which can affect normal glucose metabolism. And it has been proved that diet^[19]^ and/or physical activity^[20]^ behaviors can regulate insulin secretion and glucose metabolism. Therefore, the biological pathways linking lifestyle and *INSR* genotypes to obesity are largely overlapped, and interactions between them possibly occur on these pathways.

In our study, rs7508679 (C), rs10420008 (G), rs11883325 (C), and rs4804416 (G) alleles were found to be associated with better response to the intervention. Previous GWAS or functional studies suggest these loci are related to human metabolism. For example, Hoffmann^[21]^ et al. and Eriksson^[22]^ et al. found that carriers of the G allele (rs4804416) had a higher level of triglyceride and hypothyroidism than those non-carriers, respectively. As for rs10420008, mRNA expression data available from the Genotype-Tissue Expression (GTEx) portal suggested that carriers of GG genotype had greater hypothyroidism compared to those with AA or AG genotypes. Individuals with hypothyroidism may experience symptoms of decreased metabolic rate such as obesity. Another case-control study^[23]^ found that rs1799817 (which has a strong linkage disequilibrium with rs11883325 in the CHB population (r^2^ = 0.8)) was associated with an increased risk of nonalcoholic fatty liver disease (NAFLD). NAFLD is a metabolic stress liver disease closely related to insulin resistance, and patients usually have metabolic syndrome such as obesity. Taken together, carriers of these alleles were obesity-susceptible, and previous studies showed that such risk sub-populations tended to be more sensitive to dietary and/or physical activity interventions and thus obtained more benefits from the interventions. Nevertheless, the detailed biological mechanisms underlying the observed interactions between the *INSR* genotypes and childhood obesity interventions need further functional experiments in the future.

The observed genetic effects on the response to the intervention from baseline to 9 months did not sustain to the period after the end of the intervention (from 9 months to 31 months). The discrepancy of genetic effects between the two periods might lie in the practical barrier of keeping up healthy diet and physical activity behaviors after the end of the intervention. In contrast to our findings, a one-year lifestyle intervention study^[24]^ including 346 European children who were overweight indicated that *FTO* rs9939609 was not associated with reduction in BMI z-score during the intervention, but was associated with weight regain one year after the intervention ended. Bearing in mind that this European study did not have a comparison group and further rigorous-designed study are needed to compare genetic effects between the intervention and control groups.

Our study was unique in exploring the specific diet or physical activity behaviors that modified the effects of *INSR* gene on obesity-related indicators. If these findings were verified in future longitudinal studies, we took a further step in elucidating the specific intervention component out of the whole multifaceted intervention that might be responsible for interacting with the genetic effects. For instance, if genetic variants were interacted with the diet behavior, future personalized interventions should focus on improving diet based on individuals’ genetic background.

## Limitations and Strengths

Our study findings should be interpreted with cautions. First, the sample size of intervention studies is relatively smaller than that of observational studies due to its implementation complexity in real-world settings, and our study is not exceptional. However, our study involved multiple obesity-related indicators and showed consistent patterns with each other, making our findings less likely to be biased. Second, all study participants were Han population in Beijing, China, limiting the generalizability. Last, the mechanisms underlying the study findings need to be clarified in future functional studies.

Nevertheless, our study has several strengths. First, our study was unique in its randomized, parallel-controlled group design. The optimized study design reduced the selection and confounding biases of study findings. And this approach allowed us to analyze whether any distinct effect of *INSR* genotype on obesity is specific to the obesity intervention, by comparing obesity-related indicators between children randomized to the intervention group and those randomized to the control group. Second, our study carried out unified training for project personnel at all stages of research, including the measurement of indicators, the collection of saliva, and the extraction of genome DNA, ensuring the accuracy and reliability of analysis results.

Moreover, our study followed up children for 31 months and our observation extended to the period following the end of the intervention (from 9 months to 31 months). Last, the participants of this study included not only children who were overweight or obese but also children with normal weight, shedding light on the personalized interventions focusing on early prevention of obesity.

## Conclusion

To conclude, our study indicates that children carrying effect alleles of rs7508679, rs10420008, rs11883325, or rs4804416 may benefit more from a lifestyle intervention for obesity prevention, but the effect appeared to be attenuated in the long term. Future long-term, large-scale interventional studies are needed to study the modification role of gene on prevention of childhood obesity in other ethnicities.

## Data Availability

All data produced in the present work are contained in the manuscript

---

**Figure 1 The distribution of *INSR* gene in chromosome 19 and the distribution of selected 7 SNPs in *INSR* gene**

**Figure 2 Study flow**

## Notes

**Funding** This work was supported by the National Natural Science Foundation of China (81903343).

### Competing Interest Statement

The authors have declared no competing interest.

### Clinical Trial

NCT03665857

### Funding Statement

This work was funded by the National Natural Science Foundation of China (81903343)

### Author Declarations

Ethics committee/IRB of Peking University Institution Review Board gave ethical approval for this work (IRB00001052-20058)

