## Supplemental Material for "Effect of the *INSR* gene variants on the long-term response to a 31-month childhood obesity intervention"

**Waist-related indicators**

1. **Waist circumference, cm**
2. **Whr (waist-to-hip ratio):**

Calculated as waist circumference (cm) / hip circumference (cm)

3. **WHtR (waist-to-height ratio):**

Calculated as waist circumference (cm) / height (cm)

4. **WCadjBMI (waist circumference adjusted for BMI):**

The variable was generated according to the following three steps:

(1) Fitting a linear regression model, and obtaining the predicted value of waist circumference = β0+β1*age+β2*sex+β3*BMI;
(2) Generating residuals value (wc_res) = true value of waist circumference - the predicted value of waist circumference;
(3) Performing an inverse normal transformation on wc_res to generate WCadjBMI.
5. **WHRadjBMI (waist-to-height ratio adjusted for BMI)**

The variable was generated according to the following three steps:

(1) Fitting a linear regression model, and obtaining the predicted value of whr = β0+β1*age+β2*sex+β3*BMI;
(2) Generating residuals value (whr_res) = true value of whr - the predicted value of whr;
(3) Performing an inverse normal transformation on whr_res to generate WHRadjBMI.

**Details of variables of diet and physical activity behaviours**

| **Variables** | **Properties of variables** | **Contents of variables** |
| --- | --- | --- |
| **Diet behaviors** | | |
| Diet quality distance | continuous | **Range of score**: 0 ~ 84  **Definition**: The sum of the absolute value of all indicators (grains, vegetables, fruits, milk, beans, meat, aquatic products, eggs, beverage, snacks, water, diet type scores), which can comprehensively reflect the excess and insufficiency of the diet  0: there is neither insufficient intake nor excessive intake in the diet  1 ~ 17: moderate  18 ~ 34: a low level of dietary imbalance  35 ~ 50: a moderate level of dietary imbalance  > 50: a high level of dietary imbalance |
| Eat snacks or not | categorical | **Score**: 0 or 1  **Definition**: 1: eat snacks; 0: don’t eat snacks |
| Eat fried food or not | categorical | **Score**: 0 or 1  **Definition**: 1: eat fried food; 0: don’t eat fried food |
| **Physical activity behaviors** | | |
| Moderate-to-vigorous physical activity | continuous | **Range of score**: 0 ~ 7  **Definition**: The number of days that performing moderate-to-vigorous physical activity ≥ 1 hour per week |
| Sedentary time | continuous | **Definition**: Average daily sedentary time (min) |
| Screen time | continuous | **Definition**: Average daily screen time (min) |
