## Supplemental Tables 1-6 for "Effect of the *INSR* gene variants on the long-term response to a 31-month childhood obesity intervention"

| **Supplemental Table 1 Baseline characteristics of the study population according to *INSR* rs7508679 genotype** | | | | |
| --- | --- | --- | --- | --- |
|  | TT (n=92)^1^ | TC (n=187)^1^ | CC (n=102)^1^ | *P*^2^ |
| Age, y | 9.6 ± 0.3 | 9.5 ± 0.3 | 9.6 ± 0.3 | 0.361 |
| Sex |  |  |  | 0.260 |
| Male | 40 (43.5%) | 100 (53.5%) | 54 (52.9%) |  |
| Female | 52 (56.5%) | 87 (46.5%) | 48 (47.1%) |  |
| Group assignment |  |  |  | 0.483 |
| Intervention | 47 (51.1%) | 98 (52.4%) | 46 (45.1%) |  |
| Control | 45 (48.9%) | 89 (47.6%) | 56 (54.9%) |  |
| Height (cm) | 140.2 ± 6.5 | 139.6 ± 7.3 | 140.9 ± 6.5 | 0.560 |
| Body weight (kg) | 36.7 ± 8.9 | 35.8 ± 9.0 | 37.5 ± 9.7 | 0.697 |
| BMI (kg/m^2^) | 18.6 ± 3.7 | 18.2 ± 3.3 | 18.7 ± 3.8 | 0.916 |
| BMI Z score | 0.7 ± 1.4 | 0.6 ± 1.3 | 0.7 ± 1.6 | 0.951 |
| Waist circumference (cm) | 64.8 ± 10.3 | 63.9 ± 9.8 | 66.1 ± 10.7 | 0.588 |
| Whr | 0.84 ± 0.06 | 0.84 ± 0.05 | 0.85 ± 0.06 | 0.400 |
| WHtR | 0.46 ± 0.06 | 0.46 ± 0.06 | 0.47 ± 0.07 | 0.681 |
| WCadjBMI | -0.1 ± 1.0 | -0.03 ± 1.0 | 0.1 ± 1.0 | 0.182 |
| WHRadjBMI | -0.10 ± 1.00 | 0.01 ± 0.95 | 0.06 ± 1.10 | 0.274 |
| Body fat percentage (%) | 21.02 ± 10.11 | 19.96 ± 9.73 | 21.76 ± 10.84 | 0.636 |

^1^ Values are presented as means ± SDs for continuous variables or n (%) for categorical variables.

^2^ Linear regression model for continuous variables, and chi-square test for categorical variables.

| **Supplemental Table 2 Baseline characteristics of the study population according to *INSR* rs2115386 genotype** | | | | |
| --- | --- | --- | --- | --- |
|  | TT (n=121)^1^ | TC (n=186)^1^ | CC (n=72)^1^ | *P*^2^ |
| Age, y | 9.5 ± 0.3 | 9.6 ± 0.3 | 9.6 ± 0.3 | 0.233 |
| Sex |  |  |  | 0.318 |
| Male | 62 (51.2%) | 89 (47.9%) | 42 (58.3%) |  |
| Female | 59 (48.8%) | 97 (52.2%) | 30 (41.7%) |  |
| Group assignment |  |  |  | 0.695 |
| Intervention | 58 (47.9%) | 95 (51.1%) | 39 (54.2%) |  |
| Control | 63 (52.1%) | 91 (48.9%) | 33 (45.8%) |  |
| Height (cm) | 140.5 ± 6.2 | 139.8 ± 7.1 | 140.1 ± 7.4 | 0.386 |
| Body weight (kg) | 36.7 ± 8.6 | 36.1 ± 9.0 | 37.0 ± 10.5 | 0.789 |
| BMI (kg/m^2^) | 18.5 ± 3.4 | 18.3 ± 3.5 | 18.6 ± 3.9 | 0.947 |
| BMI Z score | 0.7 ± 1.4 | 0.6 ± 1.4 | 0.7 ± 1.5 | 0.703 |
| Waist circumference (cm) | 65.1 ± 9.8 | 64.4 ± 10.1 | 64.8 ± 11.2 | 0.574 |
| Whr | 0.85 ± 0.06 | 0.84 ± 0.05 | 0.84 ± 0.06 | 0.163 |
| WHtR | 0.46 ± 0.06 | 0.46 ± 0.06 | 0.46 ± 0.07 | 0.703 |
| WCadjBMI | 0.1 ± 0.9 | 0.02 ± 1.0 | -0.2 ± 1.0 | 0.038 |
| WHRadjBMI | 0.10 ± 1.05 | 0.02 ± 0.96 | -0.20 ± 0.99 | 0.048 |
| Body fat percentage (%) | 20.99 ± 9.82 | 20.48 ± 9.78 | 20.74 ± 11.62 | 0.743 |

^1^ Values are presented as means ± SDs for continuous variables or n (%) for categorical variables.

^2^ Linear regression model for continuous variables, and chi-square test for categorical variables.

| **Supplemental Table 3 Baseline characteristics of the study population according to *INSR* rs10420008 genotype** | | | | |
| --- | --- | --- | --- | --- |
|  | AA (n=64)^1^ | AG (n=187)^1^ | GG (n=131)^1^ | *P*^2^ |
| Age, y | 9.6 ± 0.3 | 9.5 ± 0.3 | 9.6 ± 0.3 | 0.861 |
| Sex |  |  |  | 0.692 |
| Male | 30 (46.9%) | 99 (52.9%) | 66 (50.4%) |  |
| Female | 34 (53.1%) | 88 (47.1%) | 65 (49.6%) |  |
| Group assignment |  |  |  | 0.531 |
| Intervention | 29 (45.3%) | 99 (52.9%) | 64 (48.9%) |  |
| Control | 35 (54.7%) | 88 (47.1%) | 67 (51.1%) |  |
| Height (cm) | 140.1 ± 6.2 | 140.4 ± 7.3 | 139.7 ± 6.7 | 0.566 |
| Body weight (kg) | 37.4 ± 9.6 | 36.6 ± 9.1 | 35.8 ± 9.0 | 0.252 |
| BMI (kg/m^2^) | 18.9 ± 4.0 | 18.4 ± 3.4 | 18.2 ± 3.4 | 0.199 |
| BMI Z score | 0.8 ± 1.5 | 0.7 ± 1.4 | 0.6 ± 1.5 | 0.265 |
| Waist circumference (cm) | 65.7 ± 10.7 | 64.8 ± 10.1 | 64.2 ± 10.1 | 0.322 |
| Whr | 0.85 ± 0.06 | 0.84 ± 0.06 | 0.84 ± 0.06 | 0.743 |
| WHtR | 0.47 ± 0.07 | 0.46 ± 0.06 | 0.46 ± 0.06 | 0.327 |
| WCadjBMI | -0.1 ± 1.0 | 0.02 ± 1.0 | 0.01 ± 1.0 | 0.679 |
| WHRadjBMI | -0.02 ± 0.97 | -0.05 ± 0.99 | 0.09 ± 1.03 | 0.351 |
| Body fat percentage (%) | 21.90 ± 10.71 | 20.69 ± 9.93 | 20.14 ± 10.12 | 0.276 |

^1^ Values are presented as means ± SDs for continuous variables or n (%) for categorical variables.

^2^ Linear regression model for continuous variables, and chi-square test for categorical variables.

| **Supplemental Table 4 Baseline characteristics of the study population according to *INSR* rs7248939 genotype** | | | | |
| --- | --- | --- | --- | --- |
|  | AA (n=146)^1^ | AG (n=161)^1^ | GG (n=74)^1^ | *P*^2^ |
| Age, y | 9.5 ± 0.3 | 9.6 ± 0.3 | 9.6 ± 0.3 | 0.080 |
| Sex |  |  |  | 0.609 |
| Male | 76 (51.7%) | 85 (52.8%) | 34 (46.0%) |  |
| Female | 71 (48.3%) | 76 (47.2%) | 40 (54.0%) |  |
| Group assignment |  |  |  | 0.707 |
| Intervention | 70 (47.6%) | 83 (51.5%) | 39 (52.7%) |  |
| Control | 77 (52.4%) | 78 (48.5%) | 35 (47.3%) |  |
| Height (cm) | 140.1 ± 7.1 | 139.9 ± 6.8 | 140.4 ± 6.9 | 0.866 |
| Body weight (kg) | 36.7 ± 9.5 | 36.0 ± 8.6 | 37.0 ± 9.6 | 0.857 |
| BMI (kg/m^2^) | 18.5 ± 3.6 | 18.2 ± 3.2 | 18.6 ± 3.9 | 0.929 |
| BMI Z score | 0.7 ± 1.5 | 0.6 ± 1.3 | 0.7 ± 1.6 | 0.884 |
| Waist circumference (cm) | 64.7 ± 10.6 | 64.4 ± 9.3 | 65.5 ± 11.2 | 0.703 |
| Whr | 0.84 ± 0.06 | 0.84 ± 0.05 | 0.85 ± 0.06 | 0.237 |
| WHtR | 0.46 ± 0.06 | 0.46 ± 0.06 | 0.47 ± 0.07 | 0.596 |
| WCadjBMI | -0.1 ± 0.9 | 0.02 ± 1.0 | 0.1 ± 1.1 | 0.244 |
| WHRadjBMI | -0.11 ± 0.94 | 0.05 ± 1.02 | 0.13 ± 1.06 | 0.071 |
| Body fat percentage (%) | 20.54 ± 10.57 | 20.22 ± 9.18 | 22.08 ± 11.13 | 0.440 |

^1^ Values are presented as means ± SDs for continuous variables or n (%) for categorical variables.

^2^ Linear regression model for continuous variables, and chi-square test for categorical variables.

| **Supplemental Table 5 Baseline characteristics of the study population according to *INSR* rs11671297 genotype** | | | | |
| --- | --- | --- | --- | --- |
|  | AA (n=137)^1^ | AG (n=184)^1^ | GG (n=60)^1^ | *P*^2^ |
| Age, y | 9.5 ± 0.3 | 9.6 ± 0.3 | 9.6 ± 0.3 | 0.277 |
| Sex |  |  |  | 0.283 |
| Male | 77 (56.2%) | 87 (47.3%) | 30 (50.0%) |  |
| Female | 60 (43.8%) | 97 (52.7%) | 30 (50.0%) |  |
| Group assignment |  |  |  | 0.382 |
| Intervention | 63 (46.0%) | 99 (53.8%) | 30 (50.0%) |  |
| Control | 74 (54.0%) | 85 (46.2%) | 30 (50.0%) |  |
| Height (cm) | 140.2 ± 7.4 | 140.2 ± 6.7 | 139.8 ± 6.5 | 0.611 |
| Body weight (kg) | 36.2 ± 9.5 | 36.9 ± 8.9 | 36.0 ± 9.1 | 0.969 |
| BMI (kg/m^2^) | 18.2 ± 3.6 | 18.6 ± 3.4 | 18.2 ± 3.5 | 0.727 |
| BMI Z score | 0.6 ± 1.5 | 0.8 ± 1.4 | 0.6 ± 1.4 | 0.722 |
| Waist circumference (cm) | 64.5 ± 10.8 | 65.1 ± 10.0 | 64.2 ± 9.5 | 0.877 |
| Whr | 0.84 ± 0.06 | 0.84 ± 0.06 | 0.84 ± 0.05 | 0.781 |
| WHtR | 0.46 ± 0.07 | 0.46 ± 0.06 | 0.46 ± 0.06 | 0.684 |
| WCadjBMI | 0.05 ± 1.0 | -0.03 ± 1.1 | 0.03 ± 0.90 | 0.740 |
| WHRadjBMI | 0.03 ± 0.91 | -0.04 ± 1.04 | 0.08 ± 1.06 | 0.965 |
| Body fat percentage (%) | 20.17 ± 10.92 | 21.25 ± 9.53 | 20.44 ± 10.11 | 0.693 |

^1^ Values are presented as means ± SDs for continuous variables or n (%) for categorical variables.

^2^ Linear regression model for continuous variables, and chi-square test for categorical variables.

| **Supplemental Table 6 Baseline characteristics of the study population according to *INSR* rs11883325 genotype** | | | | |
| --- | --- | --- | --- | --- |
|  | TT (n=61)^1^ | TC (n=177)^1^ | CC (n=144)^1^ | *P*^2^ |
| Age, y | 9.5 ± 0.3 | 9.6 ± 0.3 | 9.5±0.3 | 0.683 |
| Sex |  |  |  | 0.457 |
| Male | 31 (50.8%) | 96 (54.2%) | 68 (47.2%) |  |
| Female | 30 (49.2%) | 81 (45.8%) | 76 (52.8%) |  |
| Group assignment |  |  |  | 0.350 |
| Intervention | 29 (47.5%) | 96 (54.2%) | 67 (46.5%) |  |
| Control | 32 (52.5%) | 81 (45.8%) | 77 (53.5%) |  |
| Height (cm) | 141.0 ± 7.3 | 140.0 ± 6.7 | 139.8 ± 6.9 | 0.338 |
| Body weight (kg) | 37.2 ± 9.1 | 36.5 ± 9.6 | 36.2 ± 8.6 | 0.620 |
| BMI (kg/m^2^) | 18.5 ± 3.4 | 18.4 ± 3.8 | 18.3 ± 3.2 | 0.894 |
| BMI Z score | 0.7 ± 1.5 | 0.6 ± 1.5 | 0.7 ± 1.3 | 0.813 |
| Waist circumference (cm) | 65.5 ± 9.9 | 64.8 ± 10.5 | 64.2 ± 9.9 | 0.518 |
| Whr | 0.85 ± 0.05 | 0.85 ± 0.06 | 0.84 ± 0.06 | 0.254 |
| WHtR | 0.46 ± 0.06 | 0.46 ± 0.06 | 0.46 ± 0.06 | 0.680 |
| WCadjBMI | 0.2 ± 0.9 | 0.003 ± 1.0 | -0.1 ± 1.0 | 0.074 |
| WHRadjBMI | 0.10 ± 0.93 | 0.04 ± 1.03 | 0.10 ± 0.99 | 0.142 |
| Body fat percentage (%) | 20.52 ± 9.19 | 20.68 ± 10.82 | 20.82 ± 9.67 | 0.813 |

^1^ Values are presented as means ± SDs for continuous variables or n (%) for categorical variables.

^2^ Linear regression model for continuous variables, and chi-square test for categorical variables.
