## Supplemental Tables 7-9 for "Effect of the *INSR* gene variants on the long-term response to a 31-month childhood obesity intervention"

**Supplemental Table 7 Moderation by *INSR* genotype of intervention effects on change in body weight, BMI, and BMI Z-score**

| **SNP** | **Effect**  **allele** | **Other**  **allele** | **β Coefficient (95% CI) in intervention group** | **β Coefficient (95% CI) in control group** | ***P* for interaction** |
| --- | --- | --- | --- | --- | --- |
| **Effect allele effects on change in body weight** | | | | | |
| rs7508679 | C | T | 0.145 (-0.317 to 0.607) | 0.104 (- 0.482 to 0.691) | 0.974 |
| rs2115386 | C | T | -0.009 (-0.462 to 0.444) | 0.459 (-0.155 to 1.073) | 0.210 |
| rs10420008 | G | A | -0.433 (-0.905 to 0.038) | 0.401 (-0.192 to 0.995) | 0.042 |
| rs7248939 | G | A | 0.129 (-0.306 to 0.564) | 0.062 (-0.524 to 0.648) | 0.808 |
| rs11671297 | G | A | -0.048 (-0.520 to 0.424) | -0.106 (-0.722 to 0.510) | 0.831 |
| rs11883325 | C | T | -0.286 (-0.754 to 0.183) | 0.007 (-0.588 to 0.603) | 0.516 |
| rs4804416 | G | T | -0.101 (-0.548 to 0.347) | 0.019 (-0.596 to 0.633) | 0.713 |
| **Effect allele effects on change in BMI** | | | | | |
| rs7508679 | C | T | 0.102 (-0.103 to 0.307) | 0.110 (-0.131 to 0.351) | 0.895 |
| rs2115386 | C | T | -0.062 (-0.262 to 0.138) | 0.143 (-0.111 to 0.397) | 0.198 |
| rs10420008 | G | A | -0.194 (-0.402 to 0.015) | 0.192 (-0.052 to 0.436) | 0.017 |
| rs7248939 | G | A | 0.066 (-0.126 to 0.259) | 0.063 (-0.178 to 0.304) | 0.902 |
| rs11671297 | G | A | -0.047 (-0.256 to 0.161) | 0.012 (-0.242 to 0.266) | 0.813 |
| rs11883325 | C | T | -0.153 (-0.360 to 0.054) | 0.020 (-0.226 to 0.265) | 0.307 |
| rs4804416 | G | T | 0.007 (-0.191 to 0.205) | 0.070 (-0.183 to 0.323) | 0.678 |
| **Effect allele effects on change in BMI Z-score** | | | | | |
| rs7508679 | C | T | 0.047 (-0.039 to 0.132) | 0.118 (0.013 to 0.224) | 0.258 |
| rs2115386 | C | T | -0.008 (-0.092 to 0.075) | 0.027 (-0.086 to 0.140) | 0.571 |
| rs10420008 | G | A | -0.091 (-0.177 to -0.004) | 0.071 (-0.037 to 0.179) | 0.015 |
| rs7248939 | G | A | 0.042 (-0.038 to 0.122) | 0.034 (-0.072 to 0.141) | 0.791 |
| rs11671297 | G | A | -0.004 (-0.090 to 0.083) | 0.064 (-0.048 to 0.177) | 0.498 |
| rs11883325 | C | T | -0.076 (-0.161 to 0.010) | -0.008 (-0.117 to 0.101) | 0.388 |
| rs4804416 | G | T | -0.004 (-0.086 to 0.079) | 0.094 (-0.017 to 0.206) | 0.163 |

Abbreviation: BMI = body mass index; BMI Z-score = body mass index Z-score; CI = confidence interval.

**Supplemental Table 8 Moderation by *INSR* genotype of intervention effects on change in waist, whr, WHtR, WCadjBMI, and WHRadjBMI**

| **SNP** | **Effect allele** | **Other**  **allele** | **β Coefficient (95% CI) in intervention group** | **β Coefficient (95% CI) in control group** | ***P* for interaction** |
| --- | --- | --- | --- | --- | --- |
| **Effect allele effects on change in waist** | | | | | |
| rs7508679 | C | T | -0.502 (-1.158 to 0.154) | 0.546 (-0.037 to 1.129) | 0.023 |
| rs2115386 | C | T | 0.302 (-0.341 to 0.945) | 0.169 (-0.446 to 0.785) | 0.831 |
| rs10420008 | G | A | -0.718 (-1.387 to -0.049) | 0.324 (-0.271 to 0.919) | 0.017 |
| rs7248939 | G | A | -0.077 (-0.697 to 0.543) | 0.294 (-0.288 to 0.877) | 0.395 |
| rs11671297 | G | A | -0.337 (-1.006 to 0.331) | 0.142 (-0.474 to 0.759) | 0.295 |
| rs11883325 | C | T | -0.685 (-1.349 to -0.021) | 0.108 (-0.488 to 0.704) | 0.069 |
| rs4804416 | G | T | -0.598 (-1.228 to 0.032) | 0.452 (-0.160 to 1.063) | 0.022 |
| **Effect allele effects on change in whr** | | | | | |
| rs7508679 | C | T | -0.007 (-0.014 to 0.0004) | 0.007 (0.001 to 0.012) | 0.008 |
| rs2115386 | C | T | 0.002 (-0.005 to 0.009) | 0.001 (-0.004 to 0.007) | 0.933 |
| rs10420008 | G | A | -0.006 (-0.014 to 0.002) | 0.001 (-0.005 to 0.006) | 0.152 |
| rs7248939 | G | A | -0.002 (-0.009 to 0.005) | 0.003 (-0.003 to 0.008) | 0.353 |
| rs11671297 | G | A | -0.007 (-0.014 to 0.001) | 0.002 (-0.004 to 0.008) | 0.083 |
| rs11883325 | C | T | -0.008 (-0.016 to -0.001) | 0.004 (-0.002 to 0.009) | 0.008 |
| rs4804416 | G | T | -0.007 (-0.014 to 0.0004) | 0.005 (-0.001 to 0.010) | 0.025 |
| **Effect allele effects on change in WHtR** | | | | | |
| rs7508679 | C | T | -0.003 (-0.007 to 0.002) | 0.004 (-0.0002 to 0.008) | 0.041 |
| rs2115386 | C | T | 0.001 (-0.003 to 0.006) | 0.001 (-0.003 to 0.005) | 0.910 |
| rs10420008 | G | A | -0.004 (-0.009 to 0.0002) | 0.003 (-0.001 to 0.007) | 0.015 |
| rs7248939 | G | A | -0.001 (-0.005 to 0.004) | 0.003 (-0.001 to 0.007) | 0.316 |
| rs11671297 | G | A | -0.003 (-0.007 to 0.002) | 0.002 (-0.002 to 0.006) | 0.186 |
| rs11883325 | C | T | -0.005 (-0.010 to -0.0002) | 0.002 (-0.003 to 0.006) | 0.038 |
| rs4804416 | G | T | -0.003 (-0.008 to 0.001) | 0.003 (-0.001 to 0.008) | 0.040 |
| **Effect allele effects on change in WCadjBMI** | | | | | |
| rs7508679 | C | T | -0.187 (-0.337 to -0.038) | 0.100 (-0.044 to 0.243) | 0.010 |
| rs2115386 | C | T | 0.073 (-0.079 to 0.224) | -0.025 (-0.176 to 0.127) | 0.486 |
| rs10420008 | G | A | -0.094 (-0.250 to 0.063) | 0.025 (-0.121 to 0.172) | 0.292 |
| rs7248939 | G | A | -0.054 (-0.197 to 0.090) | 0.076 (-0.066 to 0.218) | 0.250 |
| rs11671297 | G | A | -0.033 (-0.190 to 0.123) | -0.004 (-0.154 to 0.145) | 0.768 |
| rs11883325 | C | T | -0.125 (-0.281 to 0.031) | 0.026 (-0.119 to 0.171) | 0.121 |
| rs4804416 | G | T | -0.153 (-0.300 to -0.006) | 0.086 (-0.064 to 0.236) | 0.035 |
| **Effect allele effects on change in WHRadjBMI** | | | | | |
| rs7508679 | C | T | -0.211 (-0.371 to -0.050) | 0.130 (-0.013 to 0.272) | 0.002 |
| rs2115386 | C | T | 0.041 (-0.121 to 0.203) | -0.016 (-0.167 to 0.135) | 0.702 |
| rs10420008 | G | A | -0.130 (-0.298 to 0.037) | -0.006 (-0.152 to 0.141) | 0.294 |
| rs7248939 | G | A | -0.090 (-0.244 to 0.064) | 0.097 (-0.043 to 0.238) | 0.099 |
| rs11671297 | G | A | -0.120 (-0.287 to 0.047) | 0.027 (-0.121 to 0.175) | 0.196 |
| rs11883325 | C | T | -0.183 (-0.351 to -0.015) | 0.081 (-0.063 to 0.225) | 0.015 |
| rs4804416 | G | T | -0.186 (-0.342 to -0.029) | 0.105 (-0.044 to 0.254) | 0.010 |

Abbreviations: CI = confidence interval; whr = waist-to-hip ratio; WHtR = waist-to-height ratio; WCadjBMI = waist circumference adjusted for BMI; WHRadjBMI = waist-to-hip ratio adjusted for BMI.

**Supplemental Table 9 Moderation by *INSR* genotype of intervention effects on change in body fat percentage**

| **SNP** | **Effect allele** | **Other**  **allele** | **β Coefficient (95% CI) in intervention group** | **β Coefficient (95% CI) in control group** | ***P* for interaction** |
| --- | --- | --- | --- | --- | --- |
| **Effect allele effects on change in body fat percentage** | | | | | |
| rs7508679 | C | T | -0.060 (-0.565 to 0.444) | 0.196 (-0.310 to 0.703) | 0.414 |
| rs2115386 | C | T | 0.044 (-0.449 to 0.538) | -0.010 (-0.538 to 0.519) | 0.894 |
| rs10420008 | G | A | -0.399 (-0.914 to 0.116) | -0.131 (-0.646 to 0.383) | 0.515 |
| rs7248939 | G | A | -0.118 (-0.592 to 0.357) | 0.228 (-0.274 to 0.730) | 0.353 |
| rs11671297 | G | A | -0.220 (-0.732 to 0.292) | -0.078 (-0.611 to 0.454) | 0.756 |
| rs11883325 | C | T | -0.443 (-0.952 to -0.065) | -0.108 (-0.625 to 0.408) | 0.362 |
| rs4804416 | G | T | -0.093 (-0.579 to 0.394) | -0.046 (-0.577 to 0.484) | 0.854 |

Abbreviations: CI = confidence interval.
