## Supplemental Tables 10-12 for "Effect of the *INSR* gene variants on the long-term response to a 31-month childhood obesity intervention"

**Supplemental Table 10 Moderation by *INSR* genotype of intervention effects on change in body weight, BMI, and BMI Z-score under dominant genetic model**

| **SNP** | **Effect**  **allele** | **Other**  **allele** | **β Coefficient (95% CI) in intervention group** | **β Coefficient (95% CI) in control group** | ***P* for interaction** |
| --- | --- | --- | --- | --- | --- |
| **Effect allele effects on change in weight** | | | | | |
| rs7508679 | C | T | 0.011 (-0.734 to 0.756) | -0.323 (-1.323 to 0.677) | 0.525 |
| rs2115386 | C | T | 0.030 (-0.669 to 0.729) | 0.225 (-0.689 to 1.139) | 0.770 |
| rs10420008 | G | A | -0.135 (-1.027 to 0.757) | 0.088 (-1.011 to 1.186) | 0.801 |
| rs7248939 | G | A | 0.427 (-0.238 to 1.092) | 0.082 (-0.799 to 0.964) | 0.488 |
| rs11671297 | G | A | -0.249 (-0.926 to 0.428) | -0.070 (-0.968 to 0.829) | 0.794 |
| rs11883325 | C | T | 0.383 (-0.504 to 1.271) | 0.328 (-0.814 to 1.470) | 0.877 |
| rs4804416 | G | T | -0.192 (-0.841 to 0.457) | -0.133 (-1.029 to 0.763) | 0.920 |
| **Effect allele effects on change in BMI** | | | | | |
| rs7508679 | C | T | 0.039 (-0.292 to 0.369) | -0.040 (-0.452 to 0.372) | 0.848 |
| rs2115386 | C | T | -0.060 (-0.368 to 0.249) | 0.073 (-0.304 to 0.450) | 0.615 |
| rs10420008 | G | A | -0.122 (-0.518 to 0.273) | 0.077 (-0.375 to 0.529) | 0.458 |
| rs7248939 | G | A | 0.187 (-0.107 to 0.481) | 0.053 (-0.308 to 0.415) | 0.520 |
| rs11671297 | G | A | -0.163 (-0.461 to 0.136) | 0.026 (-0.345 to 0.397) | 0.533 |
| rs11883325 | C | T | 0.135 (-0.257 to 0.528) | 0.148 (-0.321 to 0.617) | 0.980 |
| rs4804416 | G | T | -0.023 (-0.311 to 0.265) | -0.003 (-0.371 to 0.366) | 0.875 |
| **Effect allele effects on change in BMI Z-score** | | | | | |
| rs7508679 | C | T | 0.052 (-0.085 to 0.190) | 0.087 (-0.095 to 0.270) | 0.348 |
| rs2115386 | C | T | -0.013 (-0.141 to 0.116) | 0.030 (-0.138 to 0.197) | 0.700 |
| rs10420008 | G | A | -0.040 (-0.205 to 0.125) | 0.003 (-0.197 to 0.203) | 0.560 |
| rs7248939 | G | A | 0.100 (-0.022 to 0.222) | 0.072 (-0.088 to 0.231) | 0.702 |
| rs11671297 | G | A | -0.030 (-0.155 to 0.094) | 0.132 (-0.031 to 0.295) | 0.252 |
| rs11883325 | C | T | 0.034 (-0.130 to 0.197) | 0.064 (-0.144 to 0.272) | 0.827 |
| rs4804416 | G | T | -0.008 (-0.127 to 0.112) | 0.057 (-0.107 to 0.220) | 0.456 |

Abbreviation: BMI = body mass index; BMI Z-score = body mass index Z-score; CI = confidence interval.

**Supplemental Table 11 Moderation by *INSR* genotype of intervention effects on change in waist, whr, WHtR, WCadjBMI, and WHRadjBMI under dominant genetic model**

| **SNP** | **Effect allele** | **Other**  **allele** | **β Coefficient (95% CI) in intervention group** | **β Coefficient (95% CI) in control group** | ***P* for interaction** |
| --- | --- | --- | --- | --- | --- |
| **Effect allele effects on change in waist** | | | | | |
| rs7508679 | C | T | -0.274 (-1.336 to 0.789) | 0.383 (-0.618 to 1.384) | 0.451 |
| rs2115386 | C | T | 0.511 (-0.481 to 1.503) | -0.251 (-1.162 to 0.660) | 0.305 |
| rs10420008 | G | A | -0.870 (-2.134 to 0.393) | 0.400 (-0.697 to 1.497) | 0.116 |
| rs7248939 | G | A | -0.314 (-1.263 to 0.636) | 0.254 (-0.624 to 1.132) | 0.359 |
| rs11671297 | G | A | -0.591 (-1.551 to 0.368) | -0.071 (-0.972 to 0.830) | 0.449 |
| rs11883325 | C | T | -0.395 (-1.657 to 0.868) | 0.732 (-0.405 to 1.868) | 0.162 |
| rs4804416 | G | T | -0.636 (-1.554 to 0.283) | 0.242 (-0.655 to 1.138) | 0.176 |
| **Effect allele effects on change in whr** | | | | | |
| rs7508679 | C | T | -0.005 (-0.017 to 0.007) | 0.006 (-0.004 to 0.015) | 0.259 |
| rs2115386 | C | T | 0.001 (-0.010 to 0.012) | 0.0004 (-0.008 to 0.009) | 0.918 |
| rs10420008 | G | A | -0.005 (-0.020 to 0.009) | 0.001 (-0.009 to 0.012) | 0.433 |
| rs7248939 | G | A | -0.007 (-0.018 to 0.004) | 0.003 (-0.005 to 0.012) | 0.154 |
| rs11671297 | G | A | -0.008 (-0.019 to 0.003) | 0.0001 (-0.008 to 0.009) | 0.243 |
| rs11883325 | C | T | -0.009 (-0.023 to 0.006) | 0.009 (-0.002 to 0.020) | 0.038 |
| rs4804416 | G | T | -0.007 (-0.018 to 0.003) | 0.003 (-0.005 to 0.012) | 0.157 |
| **Effect allele effects on change in WHtR** | | | | | |
| rs7508679 | C | T | -0.001 (-0.008 to 0.006) | 0.003 (-0.004 to 0.010) | 0.384 |
| rs2115386 | C | T | 0.003 (-0.004 to 0.010) | -0.001 (-0.008 to 0.005) | 0.388 |
| rs10420008 | G | A | -0.006 (-0.015 to 0.003) | 0.004 (-0.004 to 0.011) | 0.064 |
| rs7248939 | G | A | -0.003 (-0.009 to 0.004) | 0.002 (-0.004 to 0.008) | 0.279 |
| rs11671297 | G | A | -0.005 (-0.012 to 0.002) | -0.0001 (-0.006 to 0.006) | 0.400 |
| rs11883325 | C | T | -0.003 (-0.012 to 0.005) | 0.006 (-0.002 to 0.014) | 0.102 |
| rs4804416 | G | T | -0.003 (-0.009 to 0.003) | 0.002 (-0.004 to 0.008) | 0.281 |
| **Effect allele effects on change in WCadjBMI** | | | | | |
| rs7508679 | C | T | -0.076 (-0.321 to 0.170) | 0.117 (-0.128 to 0.363) | 0.346 |
| rs2115386 | C | T | 0.067 (-0.164 to 0.298) | -0.079 (-0.302 to 0.144) | 0.445 |
| rs10420008 | G | A | -0.163 (-0.456 to 0.131) | 0.151 (-0.118 to 0.419) | 0.138 |
| rs7248939 | G | A | -0.143 (-0.362 to 0.077) | 0.034 (-0.178 to 0.247) | 0.298 |
| rs11671297 | G | A | -0.024 (-0.249 to 0.200) | -0.067 (-0.282 to 0.148) | 0.831 |
| rs11883325 | C | T | -0.230 (-0.524 to 0.063) | 0.109 (-0.171 to 0.389) | 0.074 |
| rs4804416 | G | T | -0.135 (-0.349 to 0.080) | 0.092 (-0.128 to 0.312) | 0.195 |
| **Effect allele effects on change in WHRadjBMI** | | | | | |
| rs7508679 | C | T | -0.160 (-0.424 to 0.103) | 0.169 (-0.075 to 0.413) | 0.172 |
| rs2115386 | C | T | 0.004 (-0.243 to 0.252) | -0.022 (-0.245 to 0.200) | 0.940 |
| rs10420008 | G | A | -0.181 (-0.497 to 0.135) | 0.047 (-0.221 to 0.316) | 0.280 |
| rs7248939 | G | A | -0.198 (-0.433 to 0.037) | 0.125 (-0.085 to 0.336) | 0.058 |
| rs11671297 | G | A | -0.128 (-0.369 to 0.112) | -0.028 (-0.241 to 0.185) | 0.523 |
| rs11883325 | C | T | -0.256 (-0.572 to 0.060) | 0.213 (-0.063 to 0.488) | 0.025 |
| rs4804416 | G | T | -0.191 (-0.419 to 0.038) | 0.120 (-0.099 to 0.340) | 0.069 |

Abbreviations: CI = confidence interval; whr = waist-to-hip ratio; WHtR = waist-to-height ratio; WCadjBMI = waist circumference adjusted for BMI; WHRadjBMI = waist-to-hip ratio adjusted for BMI.

**Supplemental Table 12 Moderation by *INSR* genotype of intervention effects on change in body fat percentage under dominant genetic model**

| **SNP** | **Effect allele** | **Other**  **allele** | **β Coefficient (95% CI) in intervention group** | **β Coefficient (95% CI) in control group** | ***P* for interaction** |
| --- | --- | --- | --- | --- | --- |
| **Effect allele effects on change in body fat percentage** | | | | | |
| rs7508679 | C | T | -0.096 (-0.908 to 0.716) | -0.144 (-1.009 to 0.720) | 0.742 |
| rs2115386 | C | T | 0.132 (-0.629 to 0.893) | -0.160 (-0.943 to 0.622) | 0.559 |
| rs10420008 | G | A | -0.803 (-1.770 to 0.164) | -0.711 (-1.653 to 0.230) | 0.909 |
| rs7248939 | G | A | -0.085 (-0.812 to 0.641) | 0.219 (-0.535 to 0.974) | 0.637 |
| rs11671297 | G | A | -0.607 (-1.339 to 0.125) | -0.121 (-0.900 to 0.657) | 0.401 |
| rs11883325 | C | T | -0.099 (-1.065 to 0.868) | 0.554 (-0.426 to 1.534) | 0.359 |
| rs4804416 | G | T | -0.216 (-0.922 to 0.490) | -0.242 (-1.015 to 0.531) | 0.986 |

Abbreviations: CI = confidence interval.
