## Supplemental Tables 13-15 for "Effect of the *INSR* gene variants on the long-term response to a 31-month childhood obesity intervention"

**Supplemental Table 13 Moderation by *INSR* genotype of intervention effects on change in body weight, BMI, and BMI Z-score among obese/overweight individuals**

| **SNP** | **Effect allele** | **Other**  **allele** | **β Coefficient (95% CI) in intervention group** | **β Coefficient (95% CI) in control group** | ***P* for interaction** |
| --- | --- | --- | --- | --- | --- |
| **Effect allele effects on change in body weight** | | | | | |
| rs7508679 | C | T | -0.323 (-1.261 to 0.615) | -0.919 (-2.098 to 0.260) | 0.517 |
| rs2115386 | C | T | -0.695 (-1.590 to 0.200) | 1.121 (-0.116 to 2.357) | 0.011 |
| rs10420008 | G | A | -0.520 (-1.429 to 0.390) | 0.934 (-0.295 to 2.163) | 0.053 |
| rs7248939 | G | A | 0.256 (-0.544 to 1.055) | -0.289 (-1.546 to 0.969) | 0.493 |
| rs11671297 | G | A | -0.062 (-1.034 to 0.910) | -0.753 (-2.016 to 0.511) | 0.450 |
| rs11883325 | C | T | -0.440 (-1.356 to 0.476) | 0.502 (-0.619 to 1.624) | 0.253 |
| rs4804416 | G | T | -0.211 (-1.054 to 0.632) | -0.596 (-1.826 to 0.634) | 0.563 |
| **Effect allele effects on change in BMI** | | | | | |
| rs7508679 | C | T | -0.079 (-0.489 to 0.330) | -0.246 (-0.708 to 0.216) | 0.647 |
| rs2115386 | C | T | -0.423 (-0.808 to -0.037) | 0.299 (-0.187 to 0.784) | 0.023 |
| rs10420008 | G | A | -0.261 (-0.652 to 0.129) | 0.428 (-0.059 to 0.914) | 0.012 |
| rs7248939 | G | A | 0.120 (-0.226 to 0.466) | -0.058 (-0.547 to 0.431) | 0.565 |
| rs11671297 | G | A | 0.033 (-0.390 to 0.455) | -0.300 (-0.785 to 0.184) | 0.448 |
| rs11883325 | C | T | -0.239 (-0.634 to 0.155) | 0.211 (-0.223 to 0.646) | 0.120 |
| rs4804416 | G | T | -0.043 (-0.409 to 0.324) | -0.133 (-0.613 to 0.347) | 0.867 |
| **Effect allele effects on change in BMI Z-score** | | | | | |
| rs7508679 | C | T | -0.007 (-0.162 to 0.147) | -0.046 (-0.159 to 0.068) | 0.800 |
| rs2115386 | C | T | -0.130 (-0.277 to 0.016) | 0.055 (-0.064 to 0.173) | 0.060 |
| rs10420008 | G | A | -0.100 (-0.247 to 0.047) | 0.103 (-0.016 to 0.222) | 0.012 |
| rs7248939 | G | A | 0.074 (-0.056 to 0.203) | -0.029 (-0.148 to 0.091) | 0.264 |
| rs11671297 | G | A | 0.022 (-0.137 to 0.182) | -0.069 (-0.188 to 0.050) | 0.537 |
| rs11883325 | C | T | -0.116 (-0.264 to 0.031) | 0.045 (-0.062 to 0.151) | 0.064 |
| rs4804416 | G | T | -0.017 (-0.155 to 0.121) | -0.014 (-0.132 to 0.103) | 0.844 |

Abbreviation: BMI = body mass index; BMI Z-score = body mass index Z-score; CI = confidence interval.

**Supplemental Table 14 Moderation by *INSR* genotype of intervention effects on change in waist, whr, WHtR, WCadjBMI, and WHRadjBMI among overweight/obese individuals**

| **SNP** | **Effect allele** | **Other**  **allele** | **β Coefficient (95% CI) in intervention group** | **β Coefficient (95% CI) in control group** | ***P* for interaction** |
| --- | --- | --- | --- | --- | --- |
| **Effect allele effects on change in waist** | | | | | |
| rs7508679 | C | T | -0.718 (-2.104 to 0.667) | 0.562 (-0.517 to 1.642) | 0.158 |
| rs2115386 | C | T | -0.345 (-1.672 to 0.982) | 0.634 (-0.497 to 1.764) | 0.226 |
| rs10420008 | G | A | -0.739 (-2.083 to 0.605) | 0.555 (-0.569 to 1.680) | 0.114 |
| rs7248939 | G | A | 0.538 (-0.646 to 1.721) | 0.083 (-1.059 to 1.224) | 0.583 |
| rs11671297 | G | A | -0.259 (-1.702 to 1.184) | -0.366 (-1.518 to 0.786) | 0.952 |
| rs11883325 | C | T | -1.068 (-2.439 to 0.303) | 0.652 (-0.357 to 1.660) | 0.041 |
| rs4804416 | G | T | -0.493 (-1.737 to 0.750) | 0.664 (-0.453 to 1.782) | 0.167 |
| **Effect allele effects on change in whr** | | | | | |
| rs7508679 | C | T | -0.003 (-0.018 to 0.012) | 0.004 (-0.005 to 0.014) | 0.384 |
| rs2115386 | C | T | -0.003 (-0.017 to 0.011) | 0.008 (-0.002 to 0.017) | 0.198 |
| rs10420008 | G | A | -0.003 (-0.017 to 0.011) | -0.002 (-0.012 to 0.008) | 0.774 |
| rs7248939 | G | A | 0.005 (-0.008 to 0.017) | 0.0003 (-0.010 to 0.010) | 0.522 |
| rs11671297 | G | A | -0.005 (-0.020 to 0.011) | 0.0003 (-0.010 to 0.011) | 0.404 |
| rs11883325 | C | T | -0.014 (-0.028 to 0.001) | 0.009 (0.001 to 0.018) | 0.004 |
| rs4804416 | G | T | -0.001 (-0.014 to 0.013) | 0.002 (-0.008 to 0.012) | 0.675 |
| **Effect allele effects on change in WHtR** | | | | | |
| rs7508679 | C | T | -0.004 (-0.014 to 0.006) | 0.005 (-0.003 to 0.012) | 0.155 |
| rs2115386 | C | T | -0.003 (-0.012 to 0.006) | 0.003 (-0.005 to 0.010) | 0.316 |
| rs10420008 | G | A | -0.005 (-0.014 to 0.004) | 0.005 (-0.003 to 0.013) | 0.068 |
| rs7248939 | G | A | 0.004 (-0.005 to 0.012) | 0.002 (-0.006 to 0.009) | 0.659 |
| rs11671297 | G | A | -0.001 (-0.011 to 0.009) | -0.002 (-0.010 to 0.006) | 0.893 |
| rs11883325 | C | T | -0.007 (-0.017 to 0.002) | 0.006 (-0.001 to 0.012) | 0.023 |
| rs4804416 | G | T | -0.003 (-0.011 to 0.006) | 0.006 (-0.002 to 0.013) | 0.123 |
| **Effect allele effects on change in WCadjBMI** | | | | | |
| rs7508679 | C | T | -0.030 (-0.328 to 0.269) | 0.240 (-0.003 to 0.482) | 0.187 |
| rs2115386 | C | T | 0.025 (-0.269 to 0.319) | 0.062 (-0.202 to 0.327) | 0.747 |
| rs10420008 | G | A | -0.046 (-0.341 to 0.249) | -0.014 (-0.275 to 0.247) | 0.915 |
| rs7248939 | G | A | 0.049 (-0.209 to 0.308) | 0.049 (-0.215 to 0.312) | 0.913 |
| rs11671297 | G | A | -0.004 (-0.321 to 0.312) | -0.093 (-0.355 to 0.169) | 0.773 |
| rs11883325 | C | T | -0.181 (-0.483 to 0.122) | 0.132 (-0.101 to 0.364) | 0.076 |
| rs4804416 | G | T | -0.053 (-0.323 to 0.218) | 0.166 (-0.089 to 0.422) | 0.238 |
| **Effect allele effects on change in WHRadjBMI** | | | | | |
| rs7508679 | C | T | -0.051 (-0.332 to 0.230) | 0.157 (-0.084 to 0.398) | 0.267 |
| rs2115386 | C | T | -0.052 (-0.324 to 0.221) | 0.063 (-0.195 to 0.322) | 0.492 |
| rs10420008 | G | A | -0.029 (-0.307 to 0.249) | -0.107 (-0.360 to 0.146) | 0.658 |
| rs7248939 | G | A | -0.032 (-0.277 to 0.212) | 0.056 (-0.202 to 0.315) | 0.682 |
| rs11671297 | G | A | -0.057 (-0.356 to 0.243) | 0.002 (-0.259 to 0.263) | 0.655 |
| rs11883325 | C | T | -0.202 (-0.486 to 0.081) | 0.204 (-0.023 to 0.430) | 0.023 |
| rs4804416 | G | T | -0.032 (-0.287 to 0.223) | 0.083 (-0.169 to 0.336) | 0.498 |

Abbreviations: CI = confidence interval; whr = waist-to-hip ratio; WHtR = waist-to-height ratio; WCadjBMI = waist circumference adjusted for BMI; WHRadjBMI = waist-to-hip ratio adjusted for BMI.

**Supplemental Table 15 Moderation by *INSR* genotype of intervention effects on change in body fat percentage among overweight/obese individuals**

| **SNP** | **Effect allele** | **Other**  **allele** | **β Coefficient (95% CI) in intervention group** | **β Coefficient (95% CI) in control group** | ***P* for interaction** |
| --- | --- | --- | --- | --- | --- |
| **Effect allele effects on change in body fat percentage** | | | | | |
| rs7508679 | C | T | -0.570 (-1.556 to 0.416) | 0.490 (-0.503 to 1.484) | 0.106 |
| rs2115386 | C | T | -0.687 (-1.621 to 0.246) | -0.213 (-1.260 to 0.833) | 0.537 |
| rs10420008 | G | A | -0.519 (-1.476 to 0.438) | 0.014 (-1.027 to 1.054) | 0.498 |
| rs7248939 | G | A | 0.261 (-0.587 to 1.110) | 0.143 (-0.906 to 1.193) | 0.903 |
| rs11671297 | G | A | -0.080 (-1.113 to 0.952) | -0.501 (-1.547 to 0.546) | 0.489 |
| rs11883325 | C | T | -0.701 (-1.662 to 0.259) | -0.380 (-1.324 to 0.564) | 0.595 |
| rs4804416 | G | T | -0.348 (-1.233 to 0.536) | 0.354 (-0.677 to 1.385) | 0.302 |

Abbreviations: CI = confidence interval.
