## Supplemental Tables 16-17 for "Effect of the *INSR* gene variants on the long-term response to a 31-month childhood obesity intervention"

**Supplemental Table 16 Moderation by *INSR* genotype of intervention effects on change in body weight, BMI, and BMI Z-score from 9 months to 31 months**

| **SNP** | **Effect**  **allele** | **Other**  **allele** | **β Coefficient (95% CI) in intervention group** | **β Coefficient (95% CI) in control group** | ***P* for interaction** |
| --- | --- | --- | --- | --- | --- |
| **Effect allele effects on change in body weight** | | | | | |
| rs7508679 | C | T | -0.906 (-1.771 to -0.041) | -0.005 (-0.841 to 0.831) | 0.183 |
| rs2115386 | C | T | 0.323 (-0.533 to 1.179) | -0.624 (-1.507 to 0.258) | 0.144 |
| rs10420008 | G | A | -0.167 (-1.068 to 0.735) | -0.658 (-1.503 to 0.187) | 0.522 |
| rs7248939 | G | A | 0.369 (-0.449 to 1.187) | 0.988 (0.171 to 1.805) | 0.239 |
| rs11671297 | G | A | 0.521 (-0.364 to 1.405) | 0.587 (-0.287 to 1.460) | 0.814 |
| rs11883325 | C | T | -0.135 (-1.028 to 0.759) | 0.424 (-0.419 to 1.268) | 0.318 |
| rs4804416 | G | T | -0.888 (-1.722 to -0.054) | -0.461 (-1.335 to 0.413) | 0.515 |
| **Effect allele effects on change in BMI** | | | | | |
| rs7508679 | C | T | -0.246 (-0.557 to 0.065) | 0.035 (-0.265 to 0.335) | 0.291 |
| rs2115386 | C | T | 0.155 (-0.152 to 0.462) | -0.308 (-0.623 to 0.007) | 0.039 |
| rs10420008 | G | A | -0.025 (-0.350 to 0.299) | -0.167 (-0.471 to 0.137) | 0.643 |
| rs7248939 | G | A | 0.040 (-0.255 to 0.335) | 0.241 (-0.054 to 0.536) | 0.298 |
| rs11671297 | G | A | 0.100 (-0.219 to 0.419) | 0.231 (-0.082 to 0.543) | 0.520 |
| rs11883325 | C | T | -0.016 (0.306 to -0.338) | 0.026 (-0.278 to 0.329) | 0.776 |
| rs4804416 | G | T | -0.268 (-0.569 to 0.033) | -0.097 (-0.411 to 0.218) | 0.487 |
| **Effect allele effects on change in BMI Z-score** | | | | | |
| rs7508679 | C | T | -0.077 (-0.172 to 0.018) | -0.006 (-0.104 to 0.092) | 0.454 |
| rs2115386 | C | T | 0.025 (-0.069 to 0.119) | -0.096 (-0.198 to 0.006) | 0.090 |
| rs10420008 | G | A | 0.008 (-0.091 to 0.108) | -0.074 (-0.172 to 0.025) | 0.313 |
| rs7248939 | G | A | 0.016 (-0.074 to 0.105) | 0.077 (-0.019 to 0.172) | 0.338 |
| rs11671297 | G | A | 0.015 (-0.082 to 0.112) | 0.095 (-0.006 to 0.196) | 0.258 |
| rs11883325 | C | T | 0.008 (-0.090 to 0.106) | 0.013 (-0.085 to 0.112) | 0.870 |
| rs4804416 | G | T | -0.078 (-0.170 to 0.014) | -0.047 (-0.150 to 0.055) | 0.756 |

Abbreviation: BMI = body mass index; BMI Z-score = body mass index Z-score; CI = confidence interval.

**Supplemental Table 17 Moderation by *INSR* genotype of intervention effects on change in waist, whr, WHtR, WCadjBMI, and WHRadjBMI from 9 months to 31 months**

| **SNP** | **Effect**  **allele** | **Other**  **allele** | **β Coefficient (95% CI) in intervention group** | **β Coefficient (95% CI) in control group** | ***P* for interaction** |
| --- | --- | --- | --- | --- | --- |
| **Effect allele effects on change in waist** | | | | | |
| rs7508679 | C | T | -0.085 (-0.971 to 0.801) | 0.133 (-0.756 to 1.022) | 0.183 |
| rs2115386 | C | T | -0.205 (-1.071 to 0.660) | -1.292 (-2.210 to -0.374) | 0.080 |
| rs10420008 | G | A | 0.327 (-0.587 to 1.240) | -0.726 (-1.619 to 0.167) | 0.117 |
| rs7248939 | G | A | 0.445 (-0.381 to 1.271) | 0.567 (-0.304 to 1.437) | 0.860 |
| rs11671297 | G | A | 0.414 (-0.482 to 1.309) | 0.562 (-0.360 to 1.484) | 0.804 |
| rs11883325 | C | T | 0.744 (-0.160 to 1.648) | 0.006 (-0.888 to 0.900) | 0.298 |
| rs4804416 | G | T | 0.077 (-0.776 to 0.930) | -0.085 (-1.013 to 0.844) | 0.757 |
| **Effect allele effects on change in whr** | | | | | |
| rs7508679 | C | T | 0.003 (-0.004 to 0.011) | -0.002 (-0.009 to 0.006) | 0.313 |
| rs2115386 | C | T | -0.004 (-0.011 to 0.004) | -0.008 (-0.016 to -0.001) | 0.310 |
| rs10420008 | G | A | 0.003 (-0.005 to 0.010) | -0.003 (-0.011 to 0.004) | 0.246 |
| rs7248939 | G | A | 0.003 (-0.004 to 0.010) | -0.001 (-0.008 to 0.006) | 0.434 |
| rs11671297 | G | A | 0.002 (-0.006 to 0.009) | 0.001 (-0.006 to 0.009) | 0.902 |
| rs11883325 | C | T | 0.008 (0.0002 to 0.016) | 0.004 (-0.007 to 0.008) | 0.162 |
| rs4804416 | G | T | 0.004 (-0.003 to 0.012) | -0.001 (-0.009 to 0.007) | 0.290 |
| **Effect allele effects on change in WHtR** | | | | | |
| rs7508679 | C | T | 0.001 (-0.005 to 0.007) | 0.001 (-0.004 to 0.007) | 0.982 |
| rs2115386 | C | T | -0.001 (-0.007 to 0.005) | -0.009 (-0.015 to -0.003) | 0.062 |
| rs10420008 | G | A | 0.002 (-0.004 to 0.008) | -0.003 (-0.009 to 0.002) | 0.215 |
| rs7248939 | G | A | 0.002 (-0.004 to 0.008) | 0.002 (-0.004 to 0.008) | 0.952 |
| rs11671297 | G | A | 0.001 (-0.005 to 0.008) | 0.003 (-0.003 to 0.009) | 0.692 |
| rs11883325 | C | T | 0.005 (-0.001 to 0.011) | -0.001 (-0.007 to 0.004) | 0.147 |
| rs4804416 | G | T | 0.001 (-0.004 to 0.007) | 0.0004 (-0.006 to 0.006) | 0.772 |
| **Effect allele effects on change in WCadjBMI** | | | | | |
| rs7508679 | C | T | 0.109 (-0.045 to 0.264) | 0.040 (-0.119 to 0.200) | 0.552 |
| rs2115386 | C | T | -0.184 (-0.335 to -0.034) | -0.141 (-0.308 to 0.026) | 0.742 |
| rs10420008 | G | A | 0.090 (-0.069 to 0.249) | -0.064 (-0.225 to 0.098) | 0.181 |
| rs7248939 | G | A | 0.111 (-0.032 to 0.255) | 0.013 (-0.144 to 0.170) | 0.385 |
| rs11671297 | G | A | 0.010 (-0.148 to 0.167) | -0.015 (-0.180 to 0.149) | 0.812 |
| rs11883325 | C | T | 0.098 (-0.062 to 0.259) | -0.006 (-0.165 to 0.154) | 0.335 |
| rs4804416 | G | T | 0.203 (0.057 to 0.350) | 0.055 (-0.111 to 0.222) | 0.197 |
| **Effect allele effects on change in WHRadjBMI** | | | | | |
| rs7508679 | C | T | 0.145 (-0.016 to 0.306) | -0.037 (-0.209 to 0.134) | 0.130 |
| rs2115386 | C | T | -0.145 (-0.302 to 0.012) | -0.142 (-0.321 to 0.036) | 0.998 |
| rs10420008 | G | A | 0.119 (-0.046 to 0.284) | -0.016 (-0.189 to 0.157) | 0.265 |
| rs7248939 | G | A | 0.069 (-0.081 to 0.218) | -0.064 (-0.232 to 0.104) | 0.261 |
| rs11671297 | G | A | -0.013 (-0.178 to 0.151) | -0.050 (-0.226 to 0.125) | 0.753 |
| rs11883325 | C | T | 0.113 (-0.056 to 0.282) | 0.013 (-0.158 to 0.184) | 0.400 |
| rs4804416 | G | T | 0.198 (0.045 to 0.351) | 0.007 (-0.172 to 0.186) | 0.111 |

Abbreviations: CI = confidence interval; whr = waist-to-hip ratio; WHtR = waist-to-height ratio; WCadjBMI = waist circumference adjusted for BMI; WHRadjBMI = waist-to-hip ratio adjusted for BMI.
